# Modeling of rotavirus transmission dynamics and impact of vaccination in Ghana

**DOI:** 10.1101/2020.03.12.20034801

**Authors:** Ernest O. Asare, Mohammad A. Al-Mamun, George E. Armah, Benjamin A. Lopman, Umesh D. Parashar, Fred Binka, Virginia E. Pitzer

## Abstract

**Background:** Rotavirus incidence remains relatively high in low-income countries (LICs) compared to high-income countries (HICs) after vaccine introduction. Ghana introduced monovalent rotavirus vaccine in April 2012 and despite the high coverage, vaccine performance has been modest compared to developed countries. The predictors of low vaccine effectiveness in LICs are poorly understood, and the drivers of subnational heterogeneity in rotavirus vaccine impact are unknown.

**Methods:** We used mathematical models to investigate variations in rotavirus incidence in children <5 years old in Ghana. We fit models to surveillance and case-control data from three different hospitals: Korle-Bu Teaching Hospital in Accra, Komfo Anokye Teaching Hospital in Kumasi, and War Memorial Hospital in Navrongo. The models were fitted to both pre- and post-vaccine data to estimate parameters describing the transmission rate, waning of maternal immunity, and vaccine response rate.

**Results:** The seasonal pattern and age distribution of rotavirus cases varied among the three study sites in Ghana. Our model was able to capture the spatio-temporal variations in rotavirus incidence across the three sites and showed good agreement with the age distribution of observed cases. The rotavirus transmission rate was highest in Accra and lowest in Navrongo, while the estimated duration of maternal immunity was longer (∼5 months) in Accra and Kumasi and shorter (∼3 months) in Navrongo. The proportion of infants who responded to the vaccine was estimated to be high in Accra and Kumasi and low in Navrongo.

**Conclusions:** Rotavirus vaccine impact varies within Ghana. A low vaccine response rate was estimated for Navrongo, where rotavirus is highly seasonal and incidence limited to a few months of the year. Our findings highlight the need to further explore the relationship between rotavirus seasonality, maternal immunity, and vaccine response rate to determine how they influence vaccine effectiveness and to develop strategies to improve vaccine impact.

**Highlights:**

- Marked variations in rotavirus incidence and vaccine impact within Ghana
- Similar rotavirus seasonality before and after vaccine introduction
- A shift in age distribution occurred following vaccine introduction
- The models provide satisfactory predictions of rotavirus outbreaks and vaccine impact

## Introduction

Rotavirus-associated gastroenteritis (RVGE) remains a worldwide public health problem, but the majority of severe morbidity and mortality occurs in infants in low-income countries (LICs) particularly in sub-Saharan Africa [1]. Globally, Tate et al. [2] estimated that rotavirus caused 528,000 deaths among children <5 years of age in 2000, decreasing to 215,000 deaths in 2013; sub-Saharan Africa accounted for about 47% and 56% of deaths in 2000 and 2013, respectively. Vaccination has proven to be a successful intervention strategy against severe RVGE. However, while rotavirus vaccine efficacy is high (85-99%) in high-income countries (HICs), it tends to be considerably lower (50-64%) in LICs [3-8]. Despite this disparity in vaccine efficacy, globally there has been an estimated 44% decrease in rotavirus mortality among in children <5 years old between 2005 and 2015, which can at least in part be attributed to vaccine protection [9].

In Ghana, prior to rotavirus vaccine introduction, RVGE was responsible for more than 25% of all diarrhea-caused hospitalizations and deaths occurring among children <5 years old [10-13]. In an effort to reduce the burden of RVGE-associated morbidity and mortality in the country, Ghana introduced the monovalent Rotarix vaccine (RV1) into its Expanded Program on Immunization (EPI) in April 2012, with a 2-dose schedule given at the ages 6 and 10 weeks [14]. Vaccine coverage has increased substantially from 42-56% in 2012 to above 90% in 2016, [15,16]. Early vaccine evaluations of RV1 have shown moderate and varied reduction in both rotavirus and all-cause diarrhea hospitalizations among <5-year-olds [15-17]. Additional benefits may be derived from rotavirus vaccination if strategies are identified to improve the vaccine effectiveness. Consequently, it is important to understand factors that account for this moderate vaccine impact and identify new strategies to help improve vaccine performance in Ghana as well as other LICs.

Since the introduction of rotavirus vaccine, dynamic mathematical models have been used to evaluate the impact of rotavirus vaccine on severe RVGE, rotavirus-associated deaths and all-cause diarrhea across different socio-economic settings and to assess the cost-effectiveness of the vaccine [18-23]. These modeling studies have assessed the drivers of intra-country transmission dynamics, revealed important factors that affect overall vaccine performance, and suggested strategies that could improve vaccine impact. For instance, it has been shown that failure to complete the full-dose schedule could significantly reduce vaccine impact [20].

While both modeling and observational studies indicate variation in vaccine effectiveness and rotavirus transmission dynamics within and between HICs and LICs, the predictors of low vaccine impact in LICs are poorly understood. However, factors such as season of birth [24], time of first infection [25,26], infant gut microbiome composition [27], malnutrition and co-infections [28] have been found to influence vaccine performance. Furthermore, the drivers of intra-country heterogeneity in rotavirus vaccine impact remain poorly understood. To address this, we assessed variations in the estimated vaccine response rate and other driving factors accounting for spatial variability in rotavirus incidence before and after vaccine introduction in Ghana using surveillance and case-control data from three different hospitals. A better understanding of the factors that account for this variability could help to develop strategies to improve vaccine effectiveness and impact.

## Method and Data

### Data and study area description

Data was obtained from surveillance and case-control studies conducted at three different hospitals in Ghana (Fig. 1): Korle-Bu Teaching Hospital in Accra, Komfo Anokye Teaching Hospital in Kumasi, and War Memorial Hospital in Navrongo. The sites in Accra and Kumasi are both teaching hospitals, while the one in Navrongo is a district hospital. These datasets have been described elsewhere [16]. Briefly, the surveillance data were collected from all children <2 (Navrongo) and <5 (Accra and Kumasi) years of age who presented with acute gastroenteritis. A case was defined as three or more watery stools per day and lasting less than 7 days. Stool samples were collected and tested for rotavirus using ELISA. After vaccine introduction, rotavirus immunization status was identified for all cases from the children’s immunization cards. For each hospital, the data were separated into pre- and post-vaccine periods: August 2007 to March 2012 (pre-vaccine) and April 2012 to April 2015 (post-vaccine) in Accra; January 2009 to March 2012 (pre-vaccine) and April 2012 to December 2015 (post-vaccine) in Kumasi; and July 2007 to May 2011 (pre-vaccine) and January 2013 to September 2016 (post-vaccine) in Navrongo. There were missing data from Navrongo between June 2011 and December 2012. The confirmed monthly rotavirus-positive cases from each hospital were aggregated into three age groups (0-11m, 12-23m, and 24-59m) for Accra and Kumasi and two age groups (0-11m, and 12-23m) for Navrongo.

**Fig 1.**
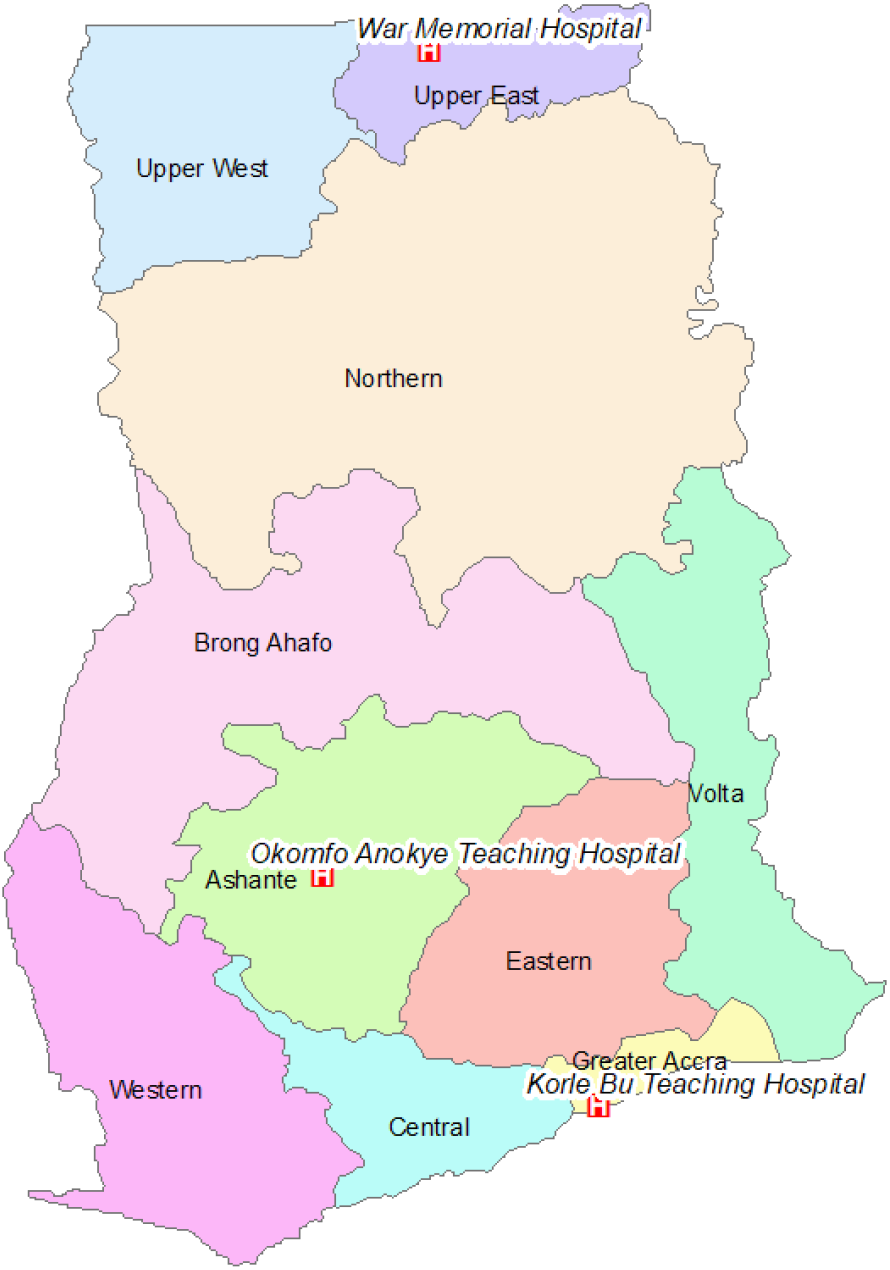
Map of Ghana showing the three hospitals.

### Model structure description

We adapted and modified an SIRS-like model developed by Pitzer et al. [18], which consists of multiple susceptible (S), infective (I), and recovered (R) compartments (Fig. 2A). Briefly, the model assumes individuals are born into the maternal class, M, at a rate equal to the annual birth rate B. All infants in this compartment are protected by maternally-acquired immunity, which gradually wanes over time at a rate *ω*_*m*_, such that individuals become fully susceptible to first infection (*S*_*0*_). Individuals in this compartment are subject to primary infection at a rate *λ*, and infected individuals are moved to the *I*_*1*_ compartment. They remain infectious for average period given by *1/γ*_*1*_, with only a fraction (*d*_*1*_) of infected individuals developing severe RVGE. Following primary infection, we assume individuals gain temporary immunity to reinfection (*R*_*1*_) that wanes at a rate *ω*, after which individuals enter the *S*_*1*_ compartment and become susceptible to secondary infection at a reduced rate *σ*_*1*_*λ*. Individuals with their second infection (*I*_*2*_) are assumed to have infectiousness lowered by a factor (*ρ*_*2*_), are less likely to develop severe RVGE (*d*_*2*_), and recover at a faster rate *γ*_*2*_ into the *R*_*2*_ compartment. Again, individuals are assumed to have temporary immunity that wanes at the same rate ω, after which they enter the partially-immune susceptible compartment (*S*_*2*_). Individuals in this compartment are subject to subsequent infections that are mostly mild or asymptomatic at a further reduced rate *σ*_*2*_*λ*. A proportion d_3_ of infected individuals (*I*_≥*3*_) could develop severe RVGE, infectiousness is reduced by a factor (*ρ*_≥*3*_), and infected individuals recover at a faster rate *γ*_*2*_. The recovered individuals are transferred to the temporary immune compartment *R*_≥*3*_ and this immunity wanes at the same rate *ω*, upon which individuals return again to the *S*_*2*_ compartment.

**Fig 2.**
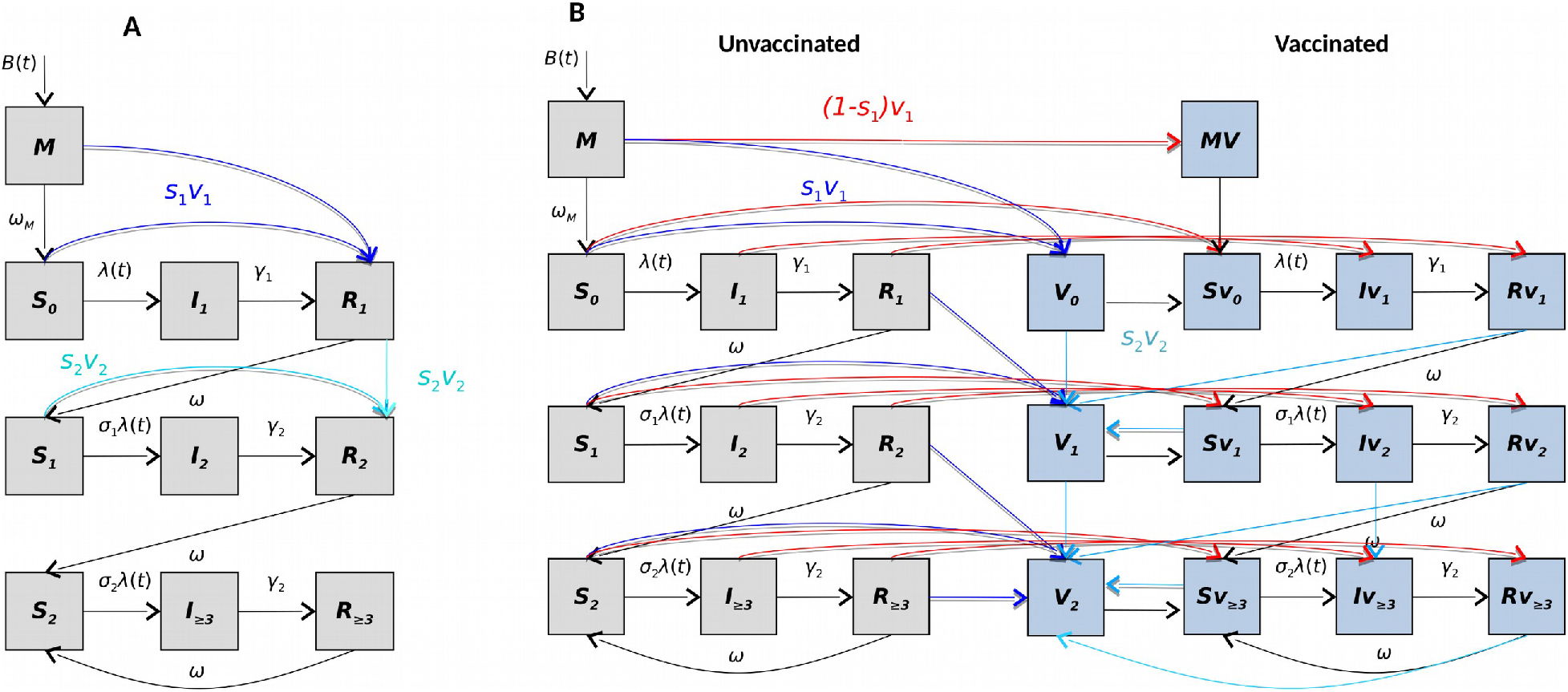
Schematic of the model structures: (A) the base model that assumes the waning of vaccine-acquired immunity is comparable to waning temporary immunity from natural infection; (B) an alternative model allows vaccine-acquired immunity to fully wane at a rate that differs from immunity derived from natural infections. The boxes indicate the various model states and lines indicate rates of movements between compartments. The red lines indicate individuals who fail to respond to the first vaccine dose, while the dark and light blue lines indicate transition of individuals who respond to the first and second doses of the vaccine, respectively.

The force of infection (transmission rate per fully susceptible individual) at time t in months, *λ*(*t*), is given by:

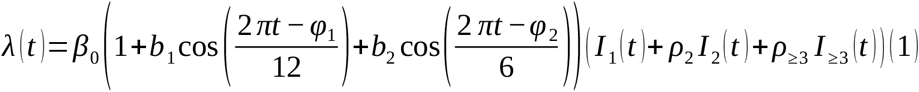

where the definitions of the parameters are given in Table 1. We assume homogeneous mixing in the population.

**Table 1:** Definition and values of fixed and estimated model parameters.

| Parameter | Symbol | Value | Reference |
| --- | --- | --- | --- |
| Basic reproductive number | $R_0$ | $=\beta_0/\gamma_1$ | Derived |
| Baseline transmission rate | $\beta_0$ | | Estimated |
| Duration of waning maternal immunity (months) | $1/\omega_m$ | | Estimated |
| Amplitude of annual seasonal forcing | $b_1$ | | Estimated |
| Annual seasonal offset (months) | $\phi_1$ | | Estimated |
| Amplitude of biannual seasonal forcing | $b_2$ | | Estimated |
| Biannual seasonal offset (months) | $\phi_2$ | | Estimated |
| Proportion of severe diarrhea cases hospitalized and reported | $h$ | | Estimated |
| Proportion of subsequent infections that are severe | $d_3$ | | Estimated |
| Proportion who respond to each vaccine dose | $s_c$ | | Estimated |
| Duration of vaccine-induced immunity (months) | $1/\omega_v$ | | Estimated |
| Duration of primary infection | $1/\gamma_1$ | 1 week | [29] |
| Duration of secondary infection | $1/\gamma_2$ | 0.5 week | [30,31] |
| Duration of temporary immunity | $1/\omega$ | 13 weeks | [32] |
| Relative risk of second infection | $\sigma_1$ | 0.62 | [33,34] |
| Relative risk of third infection | $\sigma_2$ | 0.35 | [33,34] |
| Relative infectiousness of secondary infection | $\rho_2$ | 0.5 | [35] |
| Relative infectiousness of mild/asymptomatic infections | $\rho_{\geq 3}$ | 0.1 | [35] |

Vaccination is incorporated in the model by assuming a dose of vaccine is equivalent to a natural rotavirus infection among those who respond and are immunized. Thus, individuals who are vaccinated and “respond” (i.e. seroconvert) to each vaccine dose are transferred from either the maternal immunity or susceptible or recovered compartments where vaccine is administered to the next recovered compartment. For instance, the proportion of vaccinated individuals in the M and S_0_ compartments who respond (s_c_) move directly into the R_1_ compartment after the first dose (v_1_). For the models to be consistent with the vaccine schedule in Ghana [14], we assume that infants receive their first dose upon aging into the 2-month age group and the second dose upon aging into the 3-month age group. Thus, children born on or after February 2012 were considered as vaccine-eligible for the post-vaccine simulations.

To incorporate waning of vaccine-acquired immunity in the model, we included separate compartments for vaccinated (blue) and unvaccinated (grey) individuals (Fig. 2B). The vaccinated individuals who failed to seroconvert (red lines) remain in their current state in the vaccinated compartment. For individuals who responded to the vaccine, we assumed that vaccine offers temporary protection against rotavirus infection and wanes at a rate ω_v_, which is the same for both doses. Following waning of vaccine-acquired immunity, vaccinated individuals either become susceptible to primary (after first dose) or secondary (after second dose) infection. However, individuals who responded to both doses of the vaccine remain protected and enter the next protected compartment.

### Model-fitting approach

We fit the models to the data on the age-stratified monthly number of rotavirus-positive cases from each hospital via maximum likelihood. The best-fit model parameters were estimated by minimizing the negative log-likelihood (LL) assuming that the observed number of rotavirus cases in each month and age-group was Poisson distributed with mean equal to the model-predicted number of severe RVGE cases times an estimated reporting fraction (h). We used profile likelihoods to estimate 95% confidence intervals for the model parameters of interest (R_0_, ω_m_, s_c_, and ω_v_). We adjusted for variations in h over time by multiplying it by 24-month moving average of the number of confirmed rotavirus-negative cases at each site divided by the average number of rotavirus-negative cases over the entire study period. The estimated and fixed model parameters are listed in Table 1. In order to predict the post-vaccine transmission dynamics, we made the following assumptions: (1) the proportion of vaccinated children who responded to each dose of the vaccine was equal and independent (i.e. infants who responded to the first dose were no more or less likely to respond to the second dose); (2) vaccine coverage was the same for both doses and based on the national health database; and (3) the sites had the same vaccine coverage (based on [16]).

We explored different approaches to fit the data. First, we fitted models to the pre-vaccine data only (Model A), and used the best-fit pre-vaccine parameters to predict post-vaccination cases while estimating only the proportion of individuals who responded to the vaccine and assuming infants who respond to each dose receive immunity comparable to a natural infection. Model B was similar to Model A, with the exception that we fitted models to the post-vaccination cases while estimating both the rate of waning vaccine-induced immunity and the proportion of individuals who responded to the vaccine. We then fitted models to both pre- and post-vaccination cases assuming vaccine-induced immunity is comparable to immunity from natural infection (Model C) or estimating the rate of waning vaccine-induced immunity (Model D). For Models A and C, the rate of waning of vaccine-induced immunity is assumed to be the same as the waning rate of temporary immunity from natural infection ω. We compared the fit of the four different models in each site using the Bayesian information criterion (BIC).

### Model evaluation

We further evaluated the predictive performance of the models for Navrongo by comparing to data collected between October 2016 and February 2019. This dataset from Navrongo was not used in the fitting of the model.

## Results

The transmission patterns and age distribution of reported rotavirus cases varied across the three sites (Fig. 3). In the pre-vaccination period, Accra exhibited year-round transmission with a biannual seasonal pattern; the highest number of cases typically occurred during the dry season (October-March) with occasional peaks in the wet season (May-August, e.g. in 2009) and year-to-year variation in the month of peak incidence (Fig. 3A). Rotavirus incidence in Kumasi (Fig. 3B) had a shorter peak transmission period occurring primarily during the dry season (December-February) and minor peaks during the wet season (June-August), and months with zero rotavirus cases. In Navrongo, transmission was strongly seasonal, occurring from December to March with peaks either in January or February (Fig. 3C). Outside this period, there were either zero or few isolated cases per month. Interestingly, the major peak months in Navrongo coincide with those in Kumasi. The proportion of rotavirus incidence across the age groups was similar for the three sites (Fig. 3D-F), with the majority of cases (>60%) in the 0-11-month age group. However, cases tended to be slightly younger in Navrongo and older in Kumasi.

**Fig 3.**
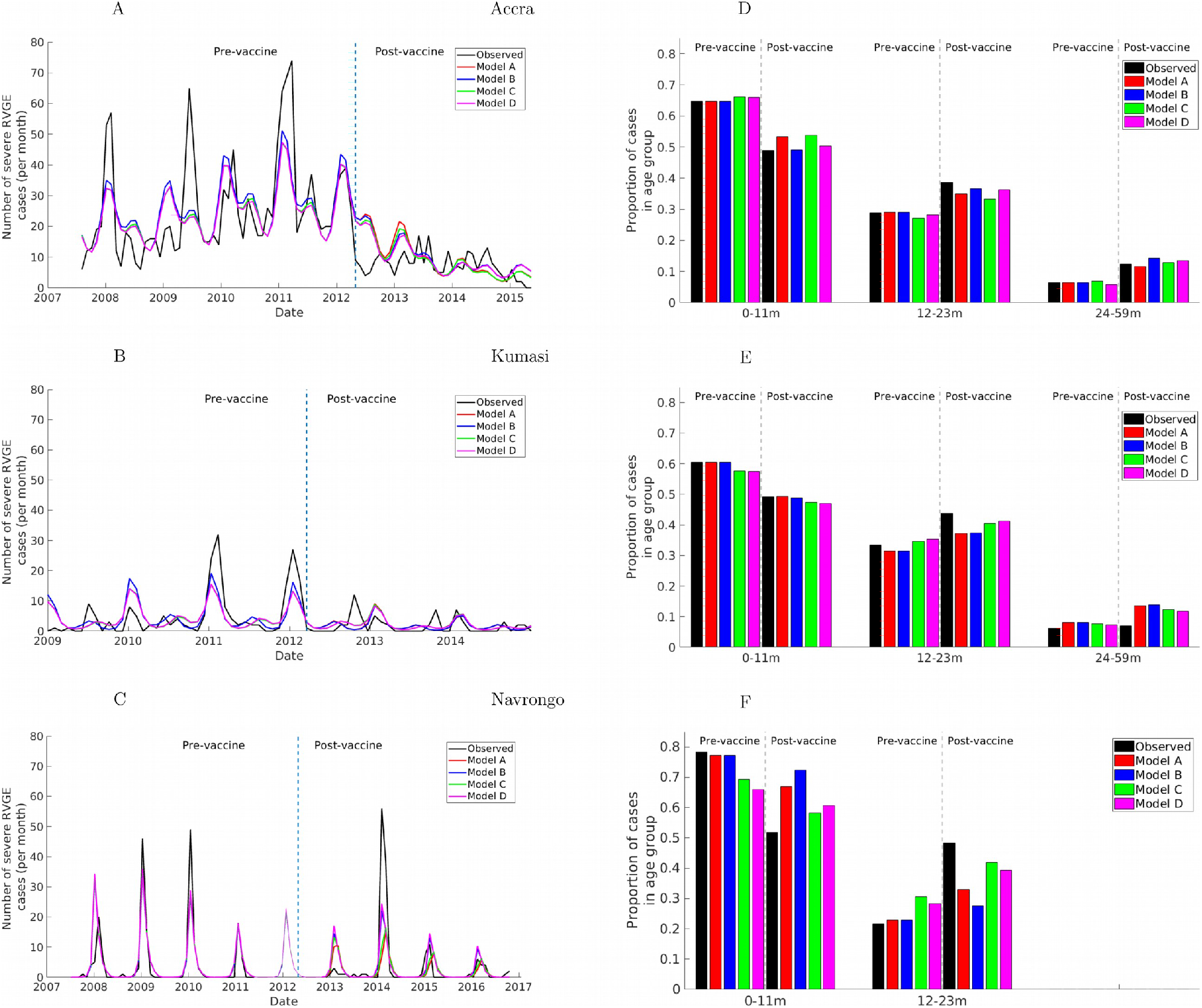
Fit of models to the observed data. (Left) Model-predicted and observed monthly incidence of severe RVGE cases among children during both pre- and post-vaccine periods. The vertical line indicates the date of vaccine introduction (April 2012). (Right) Model-predicted and observed proportion of severe RVGE in each age group before and after vaccine introduction. The grey dashed lines separate the pre- and post-vaccination distribution for each age group. The light line region of C indicates model predictions for Navrongo for the period with missing data.

Table 2 summarizes the estimated parameter values that provided the best fit to the data. The models were mostly able to capture the spatio-temporal variations in rotavirus incidence across the three sites prior to vaccine introduction, and all of the models were able to reproduce the proportion of rotavirus cases in each age group, particularly in the pre-vaccination period (Fig. 3A-C). While the models predicted the biannual transmission patterns in Accra and Kumasi, they were not able to capture the variation in the major and minor peaks in different years, in particular the major peak in June 2009 in Accra and July 2009 in Kumasi (Fig. 3A and B). In Navrongo, the models accurately predicted the observed strong seasonal transmission (Fig. 3C). However, the models tended to under- or over-estimate the peak values in all sites. All the models provided an acceptable estimate of the proportion of rotavirus cases in each age group, particularly in pre-vaccination period across the sites (Fig. 3D-F).

**Table 2:** Model estimated parameters for each study site. The symbols used in this table are explained in Table 1. Values in parentheses are 95% confidence intervals.

| **Accra** |  |  |  |  |  |  |  |  |  |  |  |
| --- | --- | --- | --- | --- | --- | --- | --- | --- | --- | --- | --- |
| **Parameter** | **$R_0$** | **$\omega_m$**<br>**months** | **$b_1$** | **$\phi_1$**<br>**months** | **$b_2$** | **$\phi_2$**<br>**months** | **$h$** | **$d_3$** | **$s_c$** | **$\omega_v$**<br>**months** | **BIC** |
| Model A | 37.861<br>(36.18-39.54) | 4.838<br>(4.44-5.22) | 0.077 | 7.203 | 0.132 | 1.037 | 0.107 | 9.54E-06 | 0.989<br>(0.84-1.0) |  | 1469.075 |
| Model B | 37.861<br>(36.18-39.54) | 4.838<br>(4.44-5.22) | 0.077 | 7.203 | 0.132 | 1.037 | 0.107 | 9.54E-06 | 0.989<br>(0.83-1.0) | 44.559<br>(26.88-55.55) | 1464.539 |
| Model C | 35.320<br>(33.98-36.7) | 4.105<br>(3.72-4.5) | 0.069 | 7.180 | 0.128 | 1.041 | 0.102 | 1.18E-05 | 0.927<br>(0.73-1.0) |  | 1463.370 |
| Model D | 38.949<br>(37.4-40.4) | 4.812<br>(4.404-5.19) | 0.074 | 7.207 | 0.119 | 1.545 | 0.101 | 2.84E-05 | 0.965<br>(0.833-1.0) | 42.366<br>(24.94-70.92) | 1459.058 |

| **Kumasi** |  |  |  |  |  |  |  |  |  |  |  |
| --- | --- | --- | --- | --- | --- | --- | --- | --- | --- | --- | --- |
| Model A | 33.661<br>(30.91-36.39) | 4.714<br>(3.492-5.87) | 0.167 | 4.127 | 0.498 | 1.055 | 0.017 | 3.09E-07 | 0.876<br>(0.55-1.0) |  | 661.636 |
| Model B | 33.661<br>(30.91-36.39) | 4.714<br>(3.492-5.87) | 0.167 | 4.127 | 0.498 | 1.055 | 0.017 | 3.09E-07 | 0.767<br>(0.43-1.0) | 5.354<br>(1.94-13.33) | 662.951 |
| Model C | 36.853<br>(34.23-39.74) | 5.698<br>(4.69-6.68) | 0.147 | 4.013 | 0.382 | 1.102 | 0.016 | 1.75E-07 | 0.856<br>(0.48-1.0) |  | 615.926 |
| Model D | 38.537<br>(35.753-41.49) | 5.987<br>(4.93-6.9) | 0.141 | 4.03 | 0.375 | 1.104 | 0.016 | 1.96E-07 | 0.836<br>(0.45-1.0) | 4.85<br>(1.25-12.35) | 617.523 |

| **Navrongo** |  |  |  |  |  |  |  |  |  |  |  |
| --- | --- | --- | --- | --- | --- | --- | --- | --- | --- | --- | --- |
| Model A | 31.529<br>(30.36-32.78) | 1.890<br>(0.39-3.18) | 0.999 | 1.050 | 6.63E-08 | 5.125 | 0.012 | 2.54E-05 | 0.299<br>(0.12-0.45) |  | 584.316 |
| Model B | 31.529<br>(30.36-32.78) | 1.890<br>(0.39-3.18) | 0.999 | 1.050 | 6.63E-08 | 5.125 | 0.012 | 2.54E-05 | 0.259<br>(0.12-0.39) | 5.806<br>(3.12-9.52) | 581.955 |
| Model C | 30.786<br>(29.89-31.73) | 3.466<br>(2.46-4.44) | 0.999 | 1.042 | 1.77E-08 | 4.142 | 0.013 | 3.46E-05 | 0.234<br>(0.07-0.44) |  | 576.094 |
| Model D | 31.212<br>(30.26-32.16) | 3.393<br>(2.4-4.32) | 0.999 | 1.043 | 4.02E-08 | 3.649 | 0.013 | 2.21E-05 | 0.230<br>(0.08-0.37) | 6.980<br>(3.38-11.91) | 573.945 |

Following vaccine introduction, seasonal transmission patterns of rotavirus cases remained relatively the same, particularly in Navrongo where dry-season epidemics continued to occur most years, with only a small peak in January 2013 (Fig. 3C). In Accra, there was a significant reduction in cases compared to the pre-vaccination period. Also, transmission remained stable throughout the year, with only slight differences between the dry and wet seasons (Fig. 3A). Similarly, in Kumasi, there was a significant decrease in cases and transmission tended to be intermittent with more months with zero cases, particularly in the wet season (Fig. 3B). Interestingly, in Accra and Kumasi, transmission tend to be relatively stable with only small differences in incidence between major and minor seasons. On the other hand, in Navrongo, a post-vaccination epidemic occurred in 2014, with incidence exceeding that of the pre-vaccination peaks; smaller epidemics also occurred in 2015 and 2016 (Fig. 3C). In terms of age distribution, there was a marked shift away from rotavirus cases occurring in the 0-11m age group towards older age groups relative to the pre-vaccination period (Fig. 3D-F). This shift in the age distribution was variable across sites, with a greater shift occurring in Navrongo.

Generally, the models were able to predict the observed reduction in the reported incidence of rotavirus in both Accra and Kumasi during the post-vaccination period. However, the models missed some noticeable peaks, and in addition, predicted a slight shift in the peak season in these two sites. Furthermore, the models overestimated the number of cases occurring in Accra during the first year following vaccine introduction. In Navrongo, while the models underestimated the post-vaccination epidemic intensity in 2014, they slightly overestimated the observed cases in 2013 and subsequent years. The models were able to capture the observed shift in the age distribution of cases to some extent, but slightly over- or under-estimated the proportion of cases in each age group relative to the observations in Accra and Kumasi (Fig. 3D and E), and were not able to fully capture the shift towards older cases during the post-vaccination period in Navrongo (Fig. 3F). The models assuming waning of vaccine-induced immunity tended to better predict the shift in cases to 12-23-month-olds, and provided a slightly better fit to the data (according to BIC).

The estimated seasonal transmission parameters (b_1_, b_2_, ϕ_1_,ϕ_2_) were similar across the models for each site, but varied between sites (Table 2). In Accra, the model predicted transmission peaks in July and January with maximum amplitude occurring in the dry season (January). The model estimated peaks in Kumasi in April and January with maximum amplitude in the dry season (January). The model was able to capture the annual transmission in Navrongo, with transmission peaking in January, whereas the observed cases peaked either in January or February.

The other important model parameters that showed the greatest variation across the three locations were the rotavirus transmission rate, duration of maternal immunity, waning of vaccine-induced immunity, and proportion who responded to the vaccine (Table 2). The estimated rotavirus transmission rate was highest in Accra (R_0_=35.3-38.9) and lowest in Navrongo (R_0_=30.8-31.5), while the mean estimated duration of maternal immunity was longer (∼5 months) in Accra and Kumasi and shorter (∼3 months) in Navrongo (Table 2). The proportion who responded to each vaccine dose was higher in Accra (s_c_=0.927-0.989) and Kumasi (s_c_=0.767-0.876), but was estimated to be very low in Navrongo (s_c_=0.230-0.299). Furthermore, the estimated duration of vaccine-induced immunity was longer in Accra (>40 months), but shorter and comparable (5-7 months) in Kumasi and Navrongo.

In Navrongo, transmission remains high and strongly seasonal 6 years following vaccine introduction, along with slightly increasing trend in intensity from 2016 (Fig. 4). There is satisfactory agreement between the models-predicted and observed post-vaccination rotavirus incidence for the evaluation period (October 2016-February 2019).

**Fig 4.**
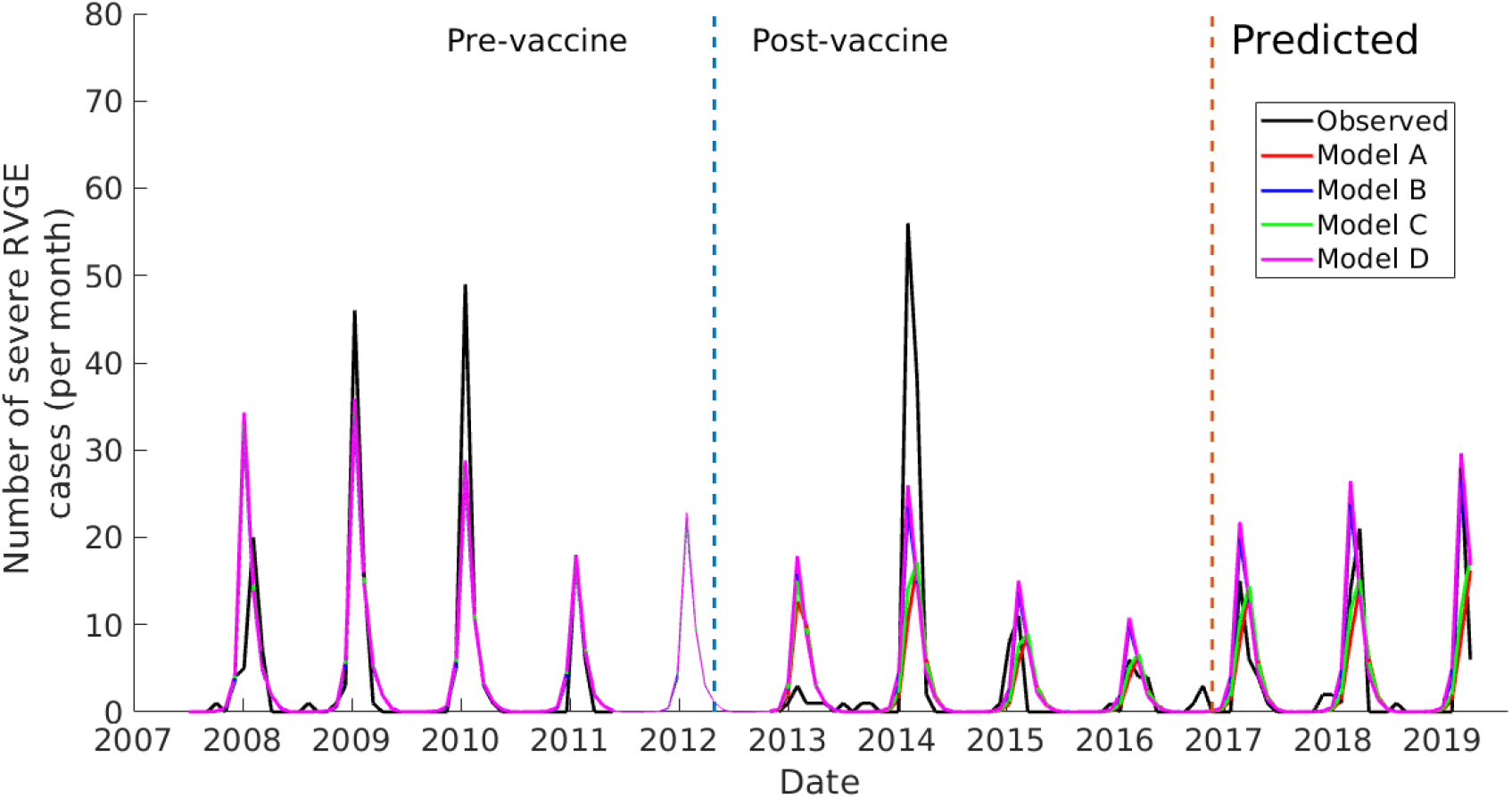
Comparison of model and observed rotavirus incidence in Navrongo showing both models fitting and evaluation periods. The brown vertical line separates the model fitting period from the evaluation period.

## Discussion

While numerous studies have documented low vaccine effectiveness in LICs relative to the HICs [21], the possible drivers accounting for this are poorly understood. Rotavirus seasonality also varies greatly between countries [36,37], but no research has attempted to investigate the potential drivers of intra-country variations in rotavirus vaccine effectiveness and its possible relationship with rotavirus seasonality. In this study, we used mathematical models to describe and quantify the variation in rotavirus transmission parameters across three sites in Ghana. Our results suggest the existence of intra-country variations in rotavirus transmission. In addition, the estimated vaccine response rate and duration of vaccine-induced immunity correlate with variation in other model parameters such as transmission seasonality, duration of maternal immunity, and transmission rates across the sites. Understanding how these different drivers of rotavirus transmission influence vaccine effectiveness could help to identify optimal strategies to improve vaccine performance across different settings, particularly in LICs.

Clear seasonal variations exist in rotavirus incidence across the three sites in Ghana (Fig. 3A-C), which are similar to those observed across countries in West Africa. Year-round rotavirus transmission with peaks during the dry season, as we and others have observed in Accra and Kumasi [13,38], has also been reported in other parts of West Africa, including Abidjan (the capital of Ivory Coast) [39] and Lome (the capital of Togo) [40]. Our model was able to reproduce this pattern, although we could not explain the increased incidence observed in both Accra and Kumasi during the 2009 wet season (Fig. 3A and B). Our models predicted a relatively stable transmission in Accra and Kumasi especially in the second year following vaccine introduction, which agrees with the observations. However, the models failed to predict the spontaneous reduction in Accra around the time vaccine was introduced, which is unlikely to be attributable to vaccination (Fig. 3A). Rotavirus vaccine was introduced in April, which coincides with the end of the rotavirus season, and only a small cohort of infants would be eligible for vaccination at that time.

In contrast, a strong seasonal pattern of rotavirus cases was observed for Navrongo for both the pre- and post-vaccine periods (Fig. 3C). Similar trends have been observed in the same study location since at least the late 1990s [41], as well as elsewhere in northern Ghana [42]. These seasonal patterns have also been observed in other West African countries. Fischer et al. [43] found the majority of cases in Guinea-Bissau occur from January to April, with few to zero cases outside these months. Another study in Enugu, Nigeria also reported this observed strong seasonal transmission with more than 90% of cases occurring between December and March over the six-year study period [44]. Similarly, in Ouagadougou (the capital of Burkina Faso), approximately 72% of acute gastroenteritis cases among children aged <5 years were rotavirus-positive during the dry season (November to May) relative to 6% rotavirus-positive during the wet season (June to October) [45]. Similar patterns were also found in the rural town of Gaoua, Burkina Faso [46]. Interestingly, Ouagadougou is an urban metropolis located about 180km from Navrongo, which is a peri-urban center located >500km from Kumasi and >800km from Accra. Despite the differences in population and economic development across study sites in West Africa, they experience a similar climate regime characterized by a prolonged dry season. While climate could be a possible driver of these dynamics, further research is required to support this hypothesis.

In addition to seasonal variations, our models also predict variation in the duration of maternal immunity, which we estimated to be shorter in Navrongo and longer in Accra and Kumasi (Table 2). The strong seasonal transmission in Navrongo may have an adverse impact on naturally acquired immunity among mothers, resulting in shorter maternal immunity duration. Consequently, infants are exposed to early rotavirus infections in Navrongo. A previous study in this same study area reported a high rotavirus incidence in infants <2 months of age [41]. Several studies have reported neonatal rotavirus infections in both LIC and HIC settings, with most infections being asymptomatic [36, 43,47]. Neonatal rotavirus infections have implications for vaccine effectiveness, as studies have shown that infants exposed to natural rotavirus infection prior to vaccination tend to have a lower seroconversion rate [25,26]. Previous rotavirus models have typically treated the duration of maternal immunity as fixed parameter [18,21-23,48]. However, our analysis reveals that duration of maternal immunity may be highly variable and should be estimated when fitting models to data from different settings.

The estimated transmission rate of rotavirus, as quantified by the basic reproductive number R_0_, was highest in Accra and lowest in Navrongo (Table 2). Despite this, the observed annual average percentage reduction in rotavirus prevalence between the pre- and post-vaccination periods was highest in Accra (74.2%) followed by Kumasi (64.4%) and Navrongo (34.9%). This differential reduction in incidence could possibly be due to variable vaccine performance, as we also estimated a higher vaccine response rate in Accra and Kumasi (77-99%) compared to Navrongo (23-30%). These results demonstrate that identifying strategies aimed at improving the vaccine response rate could reduce the burden of rotavirus even within high transmission settings.

The lower vaccine response rate seen in Navrongo could be partially explained by the shorter duration of maternal immunity and early exposure to rotavirus infection [49]. By the time the first vaccine dose is administered (6 weeks), some of the infants may have already been exposed to their first rotavirus natural infection and therefore have a lower vaccine response rate. There are studies elsewhere that support this explanation, suggesting that infants who have been infected with rotavirus before receiving vaccine were less likely to respond [25,26]. Neonatal rotavirus vaccines may provide greater protection in such a setting. Reassortant rotavirus tetravalent vaccine with the first dose administered during the neonatal period showed an efficacy of 63.1% in Navrongo [49]. Another possible explanation could be due to significant variations in the gut microbiota composition among responders and non-responder infants in Navrongo [27], thereby influencing the low vaccine response rate. However, additional studies are required to ascertain whether significant differences exist in infant gut microbiome composition across the three sites to support this hypothesis. Furthermore, we estimated a longer duration of maternal immunity and a higher vaccine response rate in Accra and Kumasi, which suggests maternal antibodies are unlikely to interfere with the vaccine response rate, as has been suggested [50,51]. This finding is supported by studies that showed withholding breastfeeding at the time of vaccine administration led to poorer vaccine effectiveness [28,52].

Comparing the pre- and post-vaccine periods provides important insights into the occurrence of rotavirus cases in various age groups (Fig. 3D-F). Both observations and our models show a similar increase in the proportion of cases in older age groups following vaccine introduction. This shift in age distribution of rotavirus cases towards >1-year-olds after vaccine introduction has been observed in both LIC and HIC settings, and for both the Rotarix and RotaTeq vaccines [23,53,54,55]. Before vaccine introduction, 90% of rotavirus hospitalizations occurred in infants aged between 3 and 12 months across five African countries [32], and in Nigeria, 66% of rotavirus cases occurred among infants aged <1 year [44]. In Belgium, Zeller et al. [54] found consistently more than more than 55% of cases in the 0-11-month age group over a seven-year period prior to vaccine introduction, decreasing to 32-43% after vaccine introduction. The almost equal proportion of infections between the 0-11m and 12-23m age groups after vaccine introduction in all the sites in Ghana is similar to observations in Burkina Faso [42]. In Malawi, there was a shift in the mean age of rotavirus caused diarrhea from 9.3 months in pre-vaccine to 11.8 months following vaccination [56].

Note that our models were able to reproduce the shift in the age distribution of cases even when we assumed the waning of vaccine-induced immunity was only partial and comparable to that of natural immunity. There are two factors that contribute to this: (1) as the vaccination program rolled out, only <1-year-olds will be directly protected by the vaccine in the first year of the program; and (2) vaccination reduces the rate of rotavirus transmission (through herd immunity), thereby delaying the time to infection among individuals who are unvaccinated, did not respond to the vaccine, or are only partially protected by vaccination. Nevertheless, waning of vaccine-induced immunity may help to explain the poor vaccine performance in Navrongo, as models in which we estimated the duration of vaccine-induced immunity provided a slightly better fit to the data and were better able to reproduce the shift in the age distribution of cases in this setting. Our findings thus highlight the need to identify vaccine strategies aimed at prolonging vaccine-induced protection in addition to the overall response rate in such settings.

The problem of low vaccine performance still persists in Navrongo 6 years following vaccine introduction. The models were able to predict the peaks of the outbreaks as well as the increasing intensity over the evaluation period. This indicates that the models provide satisfactory predictions of vaccine impact and can be used to assess the potential effectiveness of different vaccination strategies, particularly in settings with strong seasonal transmission.

## Conclusions

In summary, our results highlight the existence of marked variations in rotavirus incidence within Ghana, similar to what has been observed between countries in West Africa and other parts of the world, as well as the potential for rotavirus vaccine to reduce RVGE incidence with sustained vaccination in Ghana. However, the low vaccine impact in Navrongo presents a challenge requiring further investigation to understand how the strong seasonal transmission and shorter duration of maternal immunity might interfere with vaccine performance. While missing data in the post-vaccination period might have affected the estimated parameters for Navrongo, our models reliably predicted rotavirus incidence during the evaluation period. Our findings highlight the need to further explore the relationship between rotavirus seasonality, maternal immunity duration, and vaccine response rate and how they impact vaccine effectiveness to develop strategies to improve vaccine impact in settings with highly seasonal rotavirus transmission.

## Declaration of competing interest

VEP has received reimbursement for travel expenses from Merck to attend a Scientific Input Engagement unrelated to the subject of this paper

## Data Availability

The data that support the findings of this study are available on request from the corresponding author, EOA

## Funding

The work was supported by the National Institute of Allergy and Infectious Diseases of the National Institutes of Health [grant number R01AI112970 to V.E.P.].

